# Evaluation of Pregnant and Lactating Women’s Prescriptions for Drug-drug Interactions (DDI) And Contraindications

**DOI:** 10.1101/2022.10.17.22280909

**Authors:** Muhammad Sameer Hayat Khan

## Abstract

**Introduction:** In the pregnancy cycle and lactation period women may develop complications. Thus, special attention to pharmacotherapy is necessary, particularly to drug interactions (DDIs) and to the effect of the drugs on the foetus and the mother.

**Objective:** The aim of this study was to determine the profile of DDIs, and the risk of drugs used during pregnancy and breastfeeding among patients admitted in different hospitals.

**Methodology:** We conducted an observational and prospective study, including pregnant and breastfeeding women admitted to the different hospitals and clinics of KPK, PUNJAB and SINDH. Online databases were used to identify and classify the DDIs, and the potential risk of the drugs used during pregnancy and breastfeeding.

**Results:** On the basis of subgroups data was evaluated for pregnant and lactating women. 128 (62.0%) of the women involved in their research were pregnant women and 78 (38.0%) of them were lactating women.

In pregnant women number of interactions found were 91 (42.52%) while in lactating women number of interactions found were 123 (57.48%). Total contraindicated drugs prescribed in the prescriptions of pregnant and lactating women were 4 which was 0.48% of the total drugs prescribed from which 3 (75%) were found in prescriptions of pregnant women and 1 (15%) was found in prescription of a lactating women.

**Conclusion:** Our study demonstrated a significant number of drug- drug interactions in both pregnant and lactating mothers due to complexity in prescribed pharmacotherapy. Most of the interactions found in our prescriptions had minor to major severity level. A little percent of contraindicated drugs was also seen which could be easily substituted with safer medicine.

## INTRODUCTION

### Drug-Drug Interactions

Significant recognition of drug-drug interaction and increase in drug utilization, drug-drug interaction become the prominent talk in drug development and research. Pregnant women require special care because they are a high possibility of many obstetric complications and cause avoidable fetus harm. So a focused pharmacotherapy is required to avoid problems related to medicine that could harm both mother and child.

**“The pharmacologic or clinical response to the administration of a drug combination different from that anticipated from the known effect of the two agents when given alone**” [1]

Interaction between drugs cause considerable unwanted effect or reduce the effectiveness of therapy. Three possible outcomes of drug-drug interaction are

1. The one drug escalate the effect of other drugs
2. One drug diminishes the effect of other drugs
3. A new response may produce when drugs are given together, which is not usually seen when drugs are given separately.

In pregnancy, two souls are involved so little lethargy brings damage to both mother and fetus. To avoid these situations proper selection of medicine according to gestational period is required. Poly pharmacy and medication error are prominent causes of drug-drug interaction which increase both costs of treatment and co-morbidities.

Three types of response are seen in drug-drug interaction concerning the clinical situation.

#### The good one

In many therapeutic situations, when one drug interacting with another drug can be used to provide benefit to the patient. For example, an antidote was given after an excessive dose, to eliminate the potent effect of high dose methotrexate, leucovorin gives as an antidote.

#### The awful one

Some drug interaction consequences are pre-determined and contraindicated in practice. It is important to remember this interaction. For example, Dipyrone and enoxaparin are contraindicated in pregnancy together they cause bleeding.

#### The ugly one

Interaction with little or no therapeutic recognition has a major impact and is identified at a low level in a risk category. For example, calcium carbonate decreases the absorption of iron supplements.

### Prevalence of DDI’s

During our research different articles are evaluated and studied the percentage of (Drug-drug Interactions) is different in countries.

In Pakistan, more than one-third **(37%)** of women age 15-49 experienced at least one maternal complication or morbidity before conception. [10]

In the UK, approximately **2-3%** of all live birth are associated with a congenital anomaly. Although exogenous factors such as drugs may account for only 1-5% of this exogenous factor (**affecting <0.2% of all live births**), given that drug -drug-associated malformations are largely preventable. [11]

According to one article of Brazil, computerized prescriptions from the ICU ward of the hospital of the university were studied and data collected 305 prescriptions (200 for pregnant women and 105 for breastfeeding women) among these prescriptions 138 different drugs were prescribed, and out of which 97 drugs were involved in at least one drug interaction (70.3%) and 284 drugs have multiple drug interactions (91%).In these prescriptions, the drugs involved were (enoxaparin, Dipyrone, and hydralazine) for pregnant women, and for breastfeeding women, the drugs involved were (simethicone, Dipyrone, enoxaparin).

### Classification of Drug-Drug Interactions

DDIs classification’s main reason is that how we can predict these interactions and how to detect and overcome these interactions. So, the main classification of this is divided into 3 classes based on interactions. And these three classes are described as

1. Pharmacokinetic interactions
2. Pharmacodynamics interactions
3. Pharmaceutical interactions. [2]

### Factors Contribution to Drug-Drug Interaction

Drug-Drug Interactions are set of the complex phenomenon which cannot be predicted sometimes. Interactions can be known or they can be unknown. And unknown for the reason that each individual is different in terms of the factors given below;

1. Genetic makeup
2. Body physiology
3. Age
4. Style of living (diet, exercise)
5. Doses of drugs given
6. Time duration followed for combined therapy
7. Relativity of the time for taking doses of drugs.
8. Hydration of body
9. Co-morbidities or underlying disease conditions. [3]

### Contraindications

#### Use of contraindicated drugs during pregnancy

Contraindicated drugs in the pregnancy are the drugs that are the proportion of the drugs prescribed during pregnancy for the use which belongs to the contraindicated drugs category X, D, C, and so on according to FDA pregnancy drug categorization. In Korean research by song ET’ al, a health insurance database was used to research the use of contraindicated drug prescribing in pregnancy which was of the utmost importance. Korea has developed its list of the contraindicated drugs which should not be used by pregnant women. The use of the contraindicated drugs may lead the teratogenicity and abnormalities in the fetus and also may be fatal for the mother.

The research was performed between the years 2007 to 2011on contraindicated use of drugs in pregnancy was about 15.96% out of total prescriptions that were 355,783. And after the research and the actions taken by the authorities the contraindications in pregnancy were reduced to 11.52% later. On the whole reduction, the rate was 27.77%. Out of this greatest reduction rate was for the hormones 46.56%. The rate of reduction for the category X drugs except the hormones was 55.43%, for category D drugs it was 0.14% including the hormones. Hence, proper actions to be taken by the government for the reduction in the rate of contraindicated drug prescribing can reduce the mortality rate during pregnancy.

Most of the drugs are prescribed during pregnancy and their prevalence is increasing day by day. Most commonly prescribed drugs include, antiemetic, tranquilizers, antifibrinolytic, antihypertensives, antihistamines, analgesics, antibiotics, and antimicrobials, etc. Despite all this evidence-based research, the guidelines for drugs used during pregnancy are still not available and mainly lacking. [4]

[5] FDA has previously approved for labeling of drugs with the pregnancy categories as A, B, C, D, X. which did not ultimately prove efficacious. Hence, it developed another 3 subsections to be labeled on the drugs as per the requirements of the FDA.

1. **Pregnancy:** It includes the information of the drug’s safety during pregnancy, either the drug prescribed is safe for maternal and fetal health or not. And it also holds information that there is a specific registry that maintains the information in form of data that shows the effect of these drugs on pregnant women’s health either positive or negative effects.
2. **Lactation:** It includes the information about the use of drugs during breast-feeding or lactation period and it also refers to information that whether the drug is present in the breast milk and how much of it is present in the breast milk, and how is that drug affecting the child’s health i.e potential effects on child’s health.
3. **The reproductive potential of males and females:** It includes information about infertility, testing for pregnancy, and contraception, as it is related to the drugs.

More important is that the pregnancy and lactation will be provided with the further 3 sub-headings which are

a. Risk summary.
b. Data
c. Clinical considerations

While the third rule will not be applied to the OTC or the non-prescription drugs.

#### Rating Scale of Drug-Drug Interactions

**Table 1.** Rating scale of DDI [6]

| Categorization | Level of interaction | Therapy considerations | Cautions for use |
| --- | --- | --- | --- |
| **Category X** | **Contraindicated drugs** | Combination therapy should be avoided for such drugs. | Simultaneous use of such drugs is contraindicated in pregnancy. |
| **Category D** | **Major** | Modification of the therapy should be considered. | Medical attention is required to monitor such interactions because they may cause potential adverse effects. |
| **Category C** | **Moderate** | Drug therapy should be monitored. | The condition of a patient may get worsen due to such interactions and therapy modification becomes necessary. |
| **Category B** | **Minor** | No specific action is required in the therapy considerations. | Generally, alteration of therapy is not needed. As the interactions are not very severe and may cause an increase in severity of adverse effects. |
| **Category A** | **Unknown or no interaction** | Interaction for this category is still unknown | No interaction is known till now but research is being conducted. |

#### Why it is essential to find out DDI’s

Drug-drug interactions in the pregnancy are the curable and avoidable problems related to drugs, which causes the risk of worsening the therapeutic efficacy of drugs, responsible for adverse drug effects, causes failure of treatment, and even causes the death of the patients. That is why it is an important aspect to find out the drug-drug interactions and prevent the risk related to it and enhance the safety of medications.

Women of childbearing ages are at high risk of any drug interaction and medication errors. Which can lead to minor or even major diseases and serious complications can occur during pregnancy or after the pregnancy. As a pregnant female carrying two souls, so, it requires extra care in their treatment because sometimes a minor mistake can cause harmful effect to the mother and fetus both.

## OBJECTIVE OF THE PROJECT

The main objective of the project is to find out the potential drug-drug interactions and to designate or classify the risk of different drugs involve in lactation and pregnancy among those women, who are admitted to obstetrics and gynecology wards of the different Government and private hospitals of the KPK and Punjab its necessary to find the DDI’s in the prescriptions that we could evaluate the pharmacotherapy quality in the pregnant and lactating women. DDI’s can be fatal so it is necessary to evaluate those prescriptions and positively improve the quality of prescriptions.

### Methodology

#### Collection of Prescriptions

Total number of prescriptions collected were 231 from different hospitals and clinics of KPK, PUNJAB and SINDH (Abbottabad, Nowshera, Peshawar, Rawalpindi, Pind Dadan Khan and Thatta) in the duration of eight months i.e., from September 2020 to April 2021.

25 prescription were excluded based upon exclusion criteria (illegible) Unreadable and single-drug prescription.

#### Distribution of Cases into Sub-Group

All cases were arranged into two selected sub-groups of population.

1. Pregnant women (128 prescriptions)
2. Lactating women (78 prescriptions)

#### Distribution Of Cases On The Basis Of Age

All cases were distributed on basis of age (<20), (20-25), (26-30), (31-35), (36-40), (>40).

#### Determination and Distribution of All Prescribed Drug into Their Classes

Total prescribed drugs were 831. We analyzed the collected cases and sorted all prescribed drugs to put into respective classes of drugs like antibiotics, multivitamin and mineral supplements, analgesic and anti-emetic and so on.

#### Evaluation of Prescriptions of Both Group for Possible DDI’s And Contraindication

Prescription were evaluated by using authentic literature (Articles, Drugs Application like Medscape, Drug.com).

#### Comparison/ Distribution of DDI’s And Contraindication Found Into/ On The Basis Of

1. Sub-Groups of population selected
2. Age
3. Number Of DDIs Per Prescription
4. Number of drugs per prescription
5. Pharmacokinetic And Pharmacodynamics Mechanism

#### Data Analysis

We analyzed all the data and calculated all the percentages of number of prescriptions of both the subgroups, their age categories, potential drug-drug interactions, and contraindications in both the populations, mechanism of drug interactions, number of drugs prescribed per prescription, and number of drug classes used by using Microsoft Excel sheet in tabulated form and different graphs.

## Results

### Distribution of Cases into Sub-Groups

Based on subgroups data was evaluated for pregnant and lactating women.

128 (62.0%) of the women involved in their research were pregnant women and 78 (38.0%) of them were lactating women.

**Table 2.**
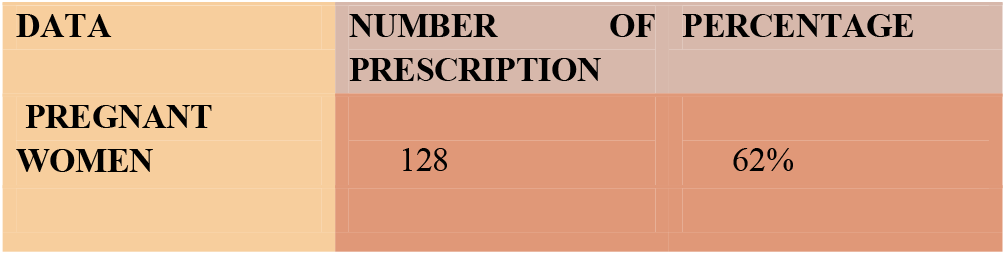
Pregnant women

**Table 3.**
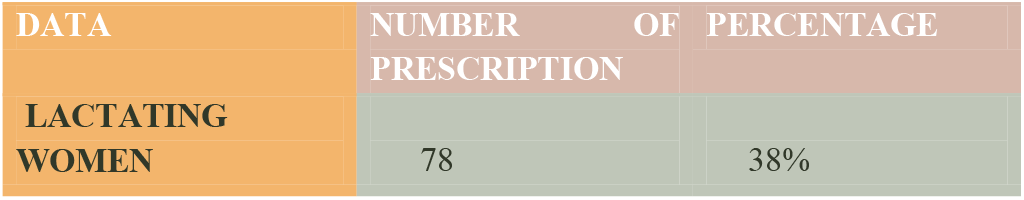
Lactating women

**Figure 1.**
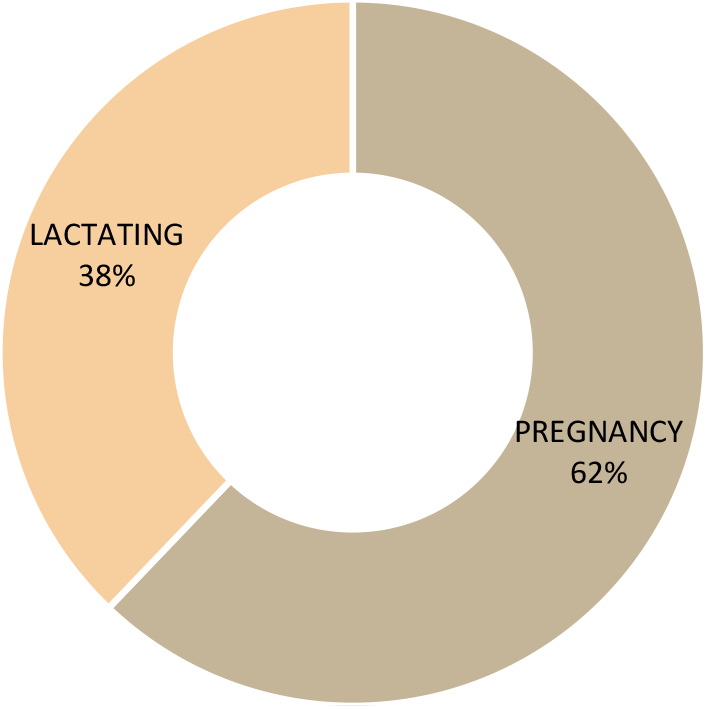
Number of Prescriptions

### Distribution of Cases On The Basis Of Age

Out of the total 206 cases evaluated, patients whose prescriptions were evaluated were categorized based on the age groups. In study 18 (8.73%) females were of age < 20, 20-25 years aged were 72 (34.95%), 26-30 years of age were 91 (44.17%), 31-35 years of aged were 21 (10.19%), 36-40 years of age were 4 (1.94%) and female of more than 40 years age was 1 (0.94%), were included.

**Table 4.** Demographic Data

| **<u>AGE</u>** | **<u>NO. OF PATIENTS</u>** | **<u>PERCENTAGE</u>** |
| --- | --- | --- |
| <20 | 18 | 8.73% |
| 20-25 | 72 | 34.95% |
| 26-30 | 91 | 44.17% |
| 31-35 | 21 | 10.19% |
| 36-40 | 4 | 1.94% |
| >40 | 1 | 0.48% |
| **TOTAL CASES (n)** | **206** |  |

**Figure 2.**
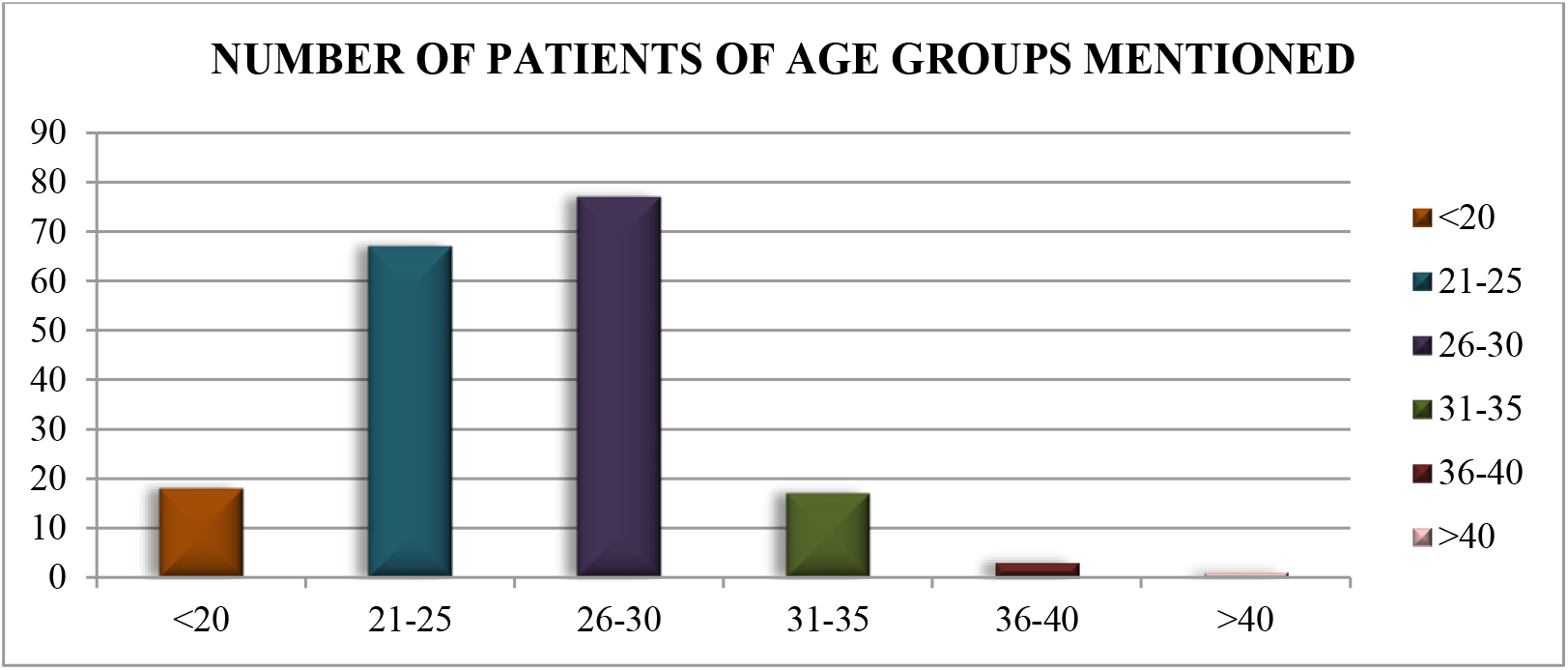
Number of patients

### Determination and Distribution of All Prescribed Drug into Their Classes

831 drugs were prescribed in 206 prescriptions including post-partum prescriptions, pre-operative prescriptions, and prescriptions of lactating or breastfeeding women. Out of these 831 drugs prescribed 213 (25.63%) were antibiotics, 69 (83.03%) were NSAIDs, 28(3.36%) were PPIs, 198(23.82%) were Vitamin, minerals, supplements, iron, and folic acid, 17(2.04%) were antifungals, 31(3.73%) were anti-fibrinolytic and anticoagulants, 7(0.84%) were antacids, 8(0.96%) were antihypertensive and anti-anginal agents, 1(0.12%) was bile acid, 37(4.45%) were anti-emetics, 14(1.68%) were herbal drugs, 14(1.68%) were corticosteroids, 9(1.08%) were antispasmodic, 28(3.36%) were hormones and enzymes, 81(9.74%) were analgesics and antipyretics, 8(0.96%) opioids and benzodiazepine and 68(8.18%) were miscellaneous including intravenous infusions and drips, etc.

**Table 5.** Distribution of Drugs

| CLASSES OF DRUGS | NUMBERS FOUND IN PRESCRIPTIONS |
| --- | --- |
| Antibiotic | 213 (25.63%) |
| NSAID's | 69 (8.03%) |
| PPI | 28 (3.36%) |
| Vit, minerals, supplements,iron,folic acid | 198 (23.82%) |
| Antifungal | 17 (2.04%) |
| Anti-fibrionolytic and anticoagulants | 31 (3.73%) |
| Antacid | 7 (0.84%) |
| Antihypertensive and anti angina | 8 (0.96%) |
| Bile acid | 1 (0.12%) |
| Anti-emetic | 37 (4.45%) |
| Herbal | 14 (1.68%) |
| Corticosteroids | 14 (1.68%) |
| Anti spasmodic | 9 (1.08%) |
| Hormones and enzymes | 28 (3.36%) |
| Analgesics and antipyretics | 81 (9.74%) |
| Opioids and benzodiazipines | 8 (0.96%) |
| Miscellaneous | 68 (8.18%) |

**Figure 3.**
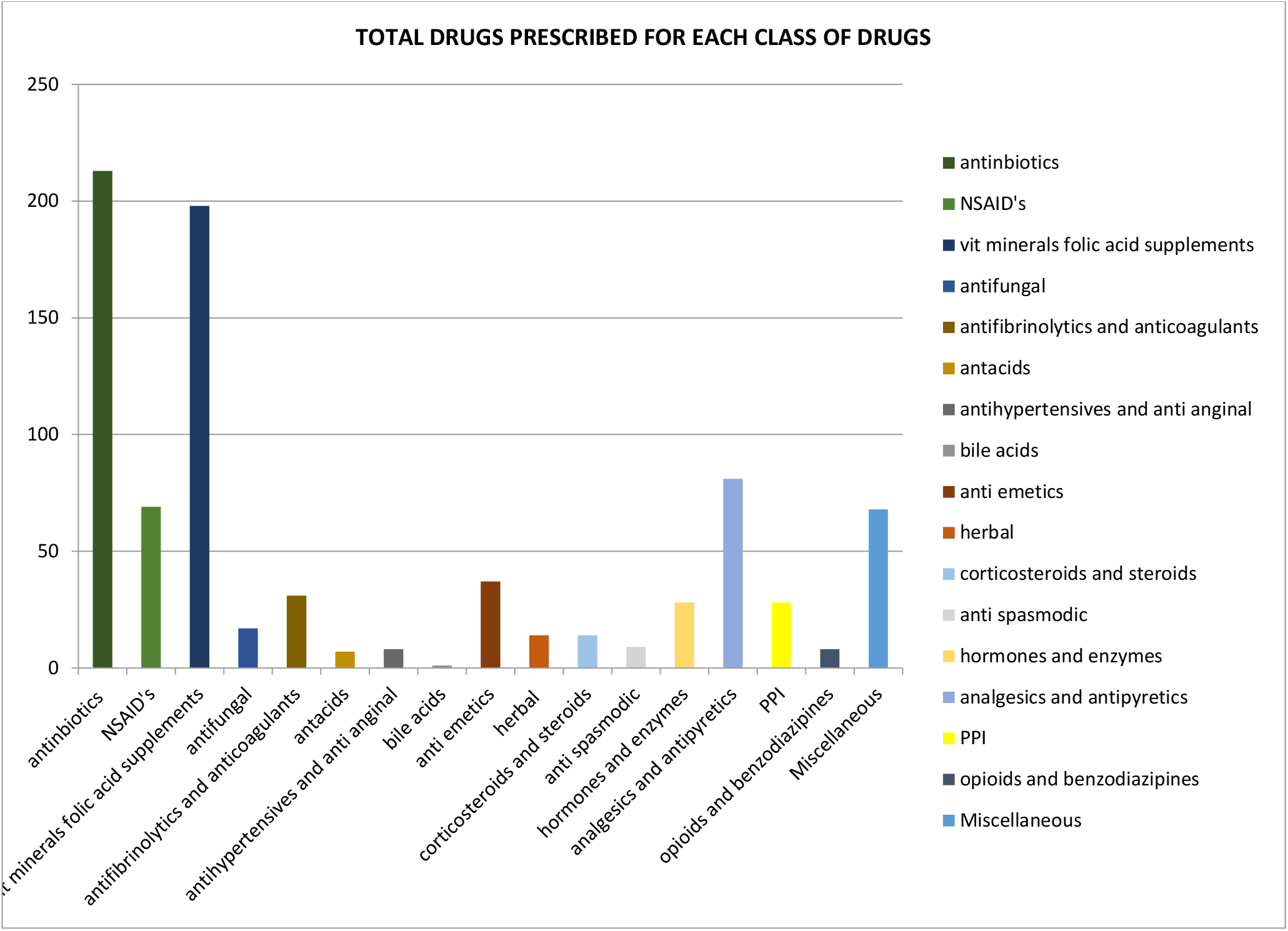
Total drugs prescribed

### Number of Drugs Prescribed Per Prescriptions

On the basis of prescribed drugs, cases were divided into 7 categories i.e. Cases in which 2, 3, 4, 5, 6, 7 and >7 drugs are prescribed. Total cases were 206. Out of them, in 23(11.16%) cases 2 drugs were prescribed, in 49(23.78%) cases 3 drugs were prescribed, 4 drugs in 36(17.47%) cases, 5 drugs in 33(16.01%) cases, 6 drugs in 35(16.99%) cases, 7 drugs in 17(8.25%) cases and more than 7 drugs were prescribed in 13(6.31%) cases.

**Table 6.** number of drug prescribed per prescription

| Number of drug prescribed | Number of prescriptions | Percentage |
| --- | --- | --- |
| 2 | 23 | 11.16% |
| 3 | 49 | 23.78% |
| 4 | 36 | 17.47% |
| 5 | 33 | 16.01% |
| 6 | 35 | 16.99% |
| 7 | 17 | 8.25% |
| >7 | 13 | 6.31% |
| **n** | **206** |  |

**Figure 4.**
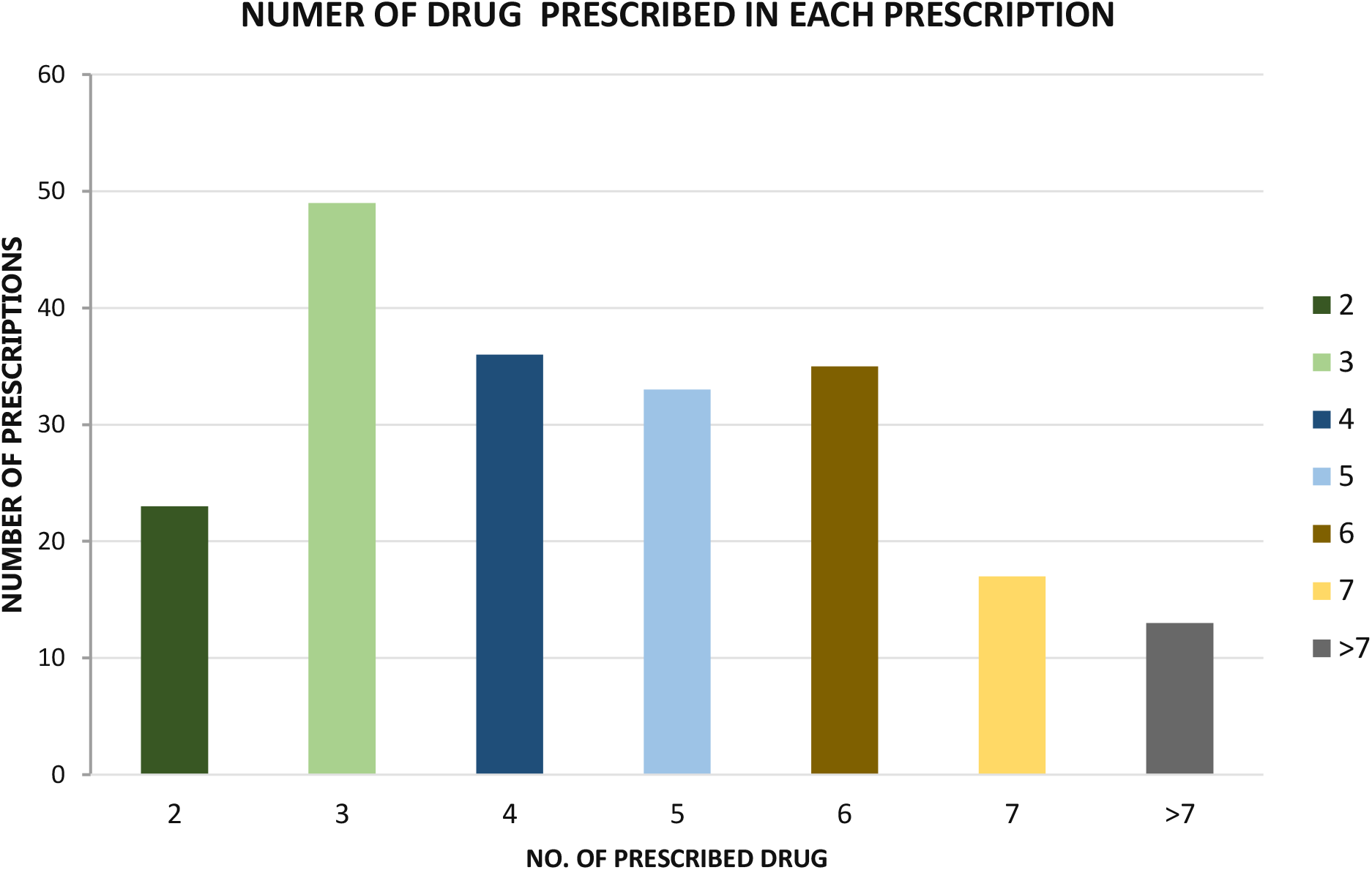
number of prescribed drugs

### Evaluation of Prescription of Both Group for Possible DDI’s and Contraindication

Pregnant and lactating women’s prescription were evaluated for possible Drug-Drug Interactions. In pregnant women number of interactions found was 91 (42.52% of total DDIs) while in lactating women number of DDIs found were 123 (57.48% of total DDIs). Total contraindicated drugs prescribed in the prescriptions of pregnant and lactating women were 4 which was 0.48% of the total drugs prescribed from which 3 (75% of total contraindications found) were found in prescriptions of pregnant women and 1 (25% of total contraindications found) was found in the prescription of a lactating woman.

**Table 7.** Population Group

| GROUP | NUMBER OF DDI'S | PERCENTAGE OF TOTAL DDIs |
| --- | --- | --- |
| PREGNANT WOMEN | 91 | 42.52% |
| LACTATING MOTHERS | 123 | 57.48% |

**Figure 5.**
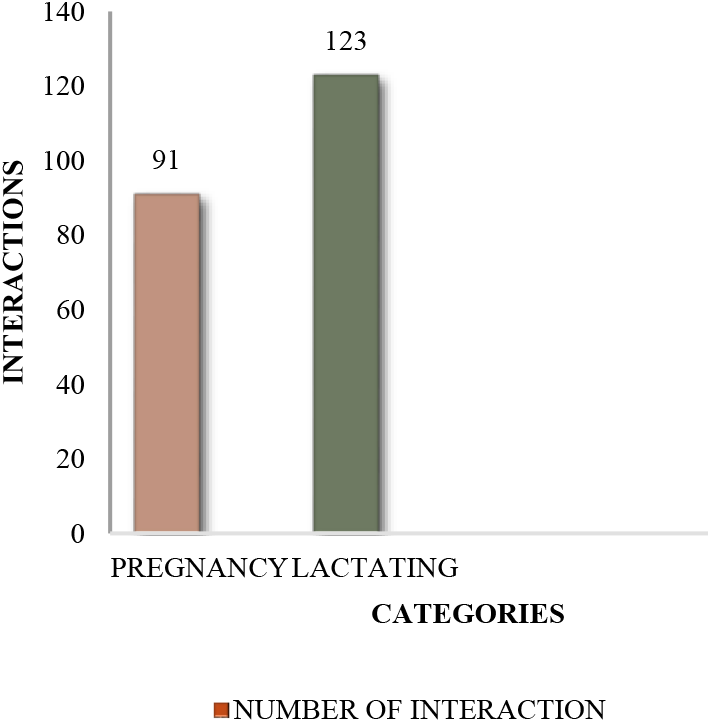
Number of DDI’S

**Figure 6.**
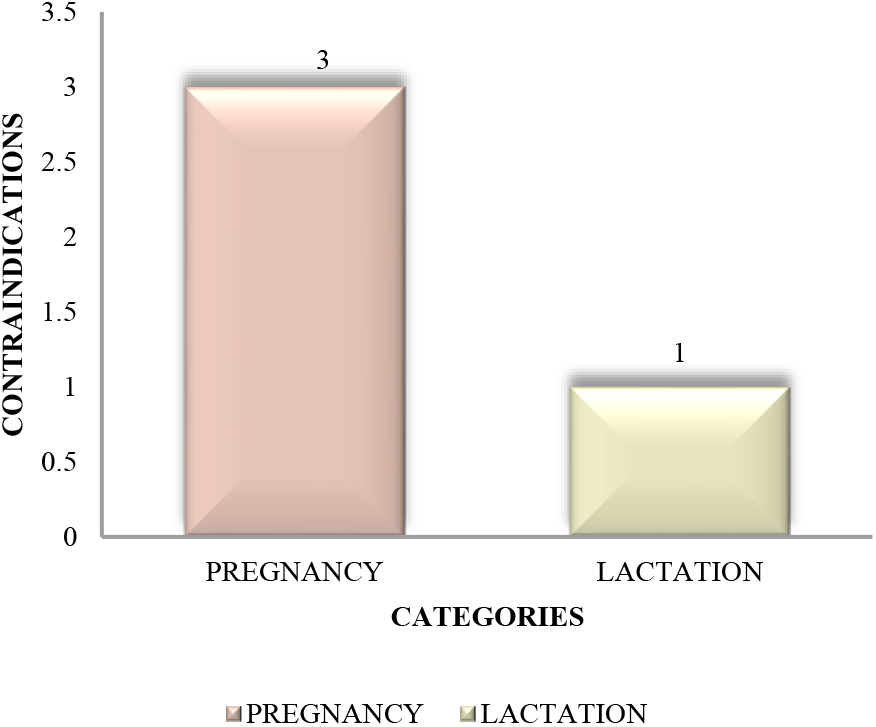
Contraindications

### Category Based DDI’s And Contraindication Distribution In pregnant Women

In pregnant women, out of 91 total interactions found based on FDA risk categorization, 27 (29.67%) were of minor severity, 33 (36.26%) were of major severity, 35 (38.46%) were of moderate severity and 3 (3.30%) were belonging to Category X of FDA risk categorization and hence, placed in contraindicated drugs.

**Table 8.** category based distribution

| **CATEGORY OF DDIs**<br>**FOUND** | **<u>NUMBER OF DDIs</u>** | **<u>PERCENTAGE OF TOTAL DDIs</u>**<br>**<u>FOUND IN PREGNANT</u>**<br>**<u>WOMEN'S PRESCRIPTIONS</u>** |
| --- | --- | --- |
| Minor | **27** | 29.67% |
| Major | **33** | 36.26% % |
| Moderate | **35** | 38.46% |
| Contraindicated | **3** | 3.30% |
| **N** | **91** |  |

**Figure 7.**
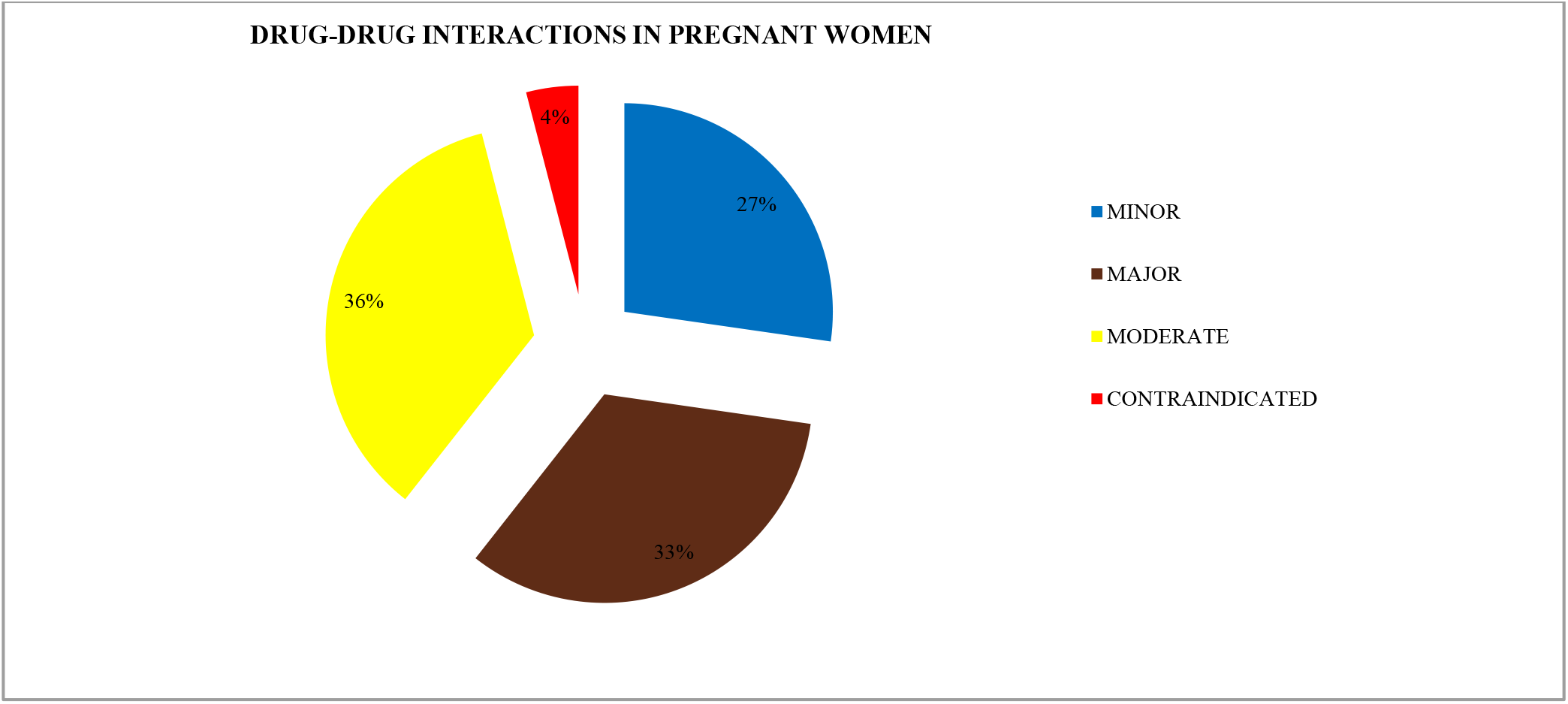
DDI’s and contraindication in pregnant women

### Category Based DDI’s and Contraindication Distribution in Lactating Women

Out of 123 interactions found in the lactating women, 48 (39.02%) were of minor severity, 42 (34.15%) were of major severity, 31 (25.20%) were of moderate severity and 1(0.81%) as belonging to category X of the FDA risk categorization i.e. contraindicated drug.

**Table 9.** category based distribution

| CATEGORY OF DDIs FOUND | <u>NUMBER OF DDIs</u> | <u>PERCENTAGE OF TOTAL DDIs FOUND IN LACTATING WOMEN'S PRESCRIPTIONS</u> |
| --- | --- | --- |
| Minor | 48 | 39.02% |
| Major | 42 | 34.15% |
| Moderate | 31 | 25.20% |
| Contraindicated | 1 | 0.81% |
| **N** | **123** |  |

**Figure 8.**
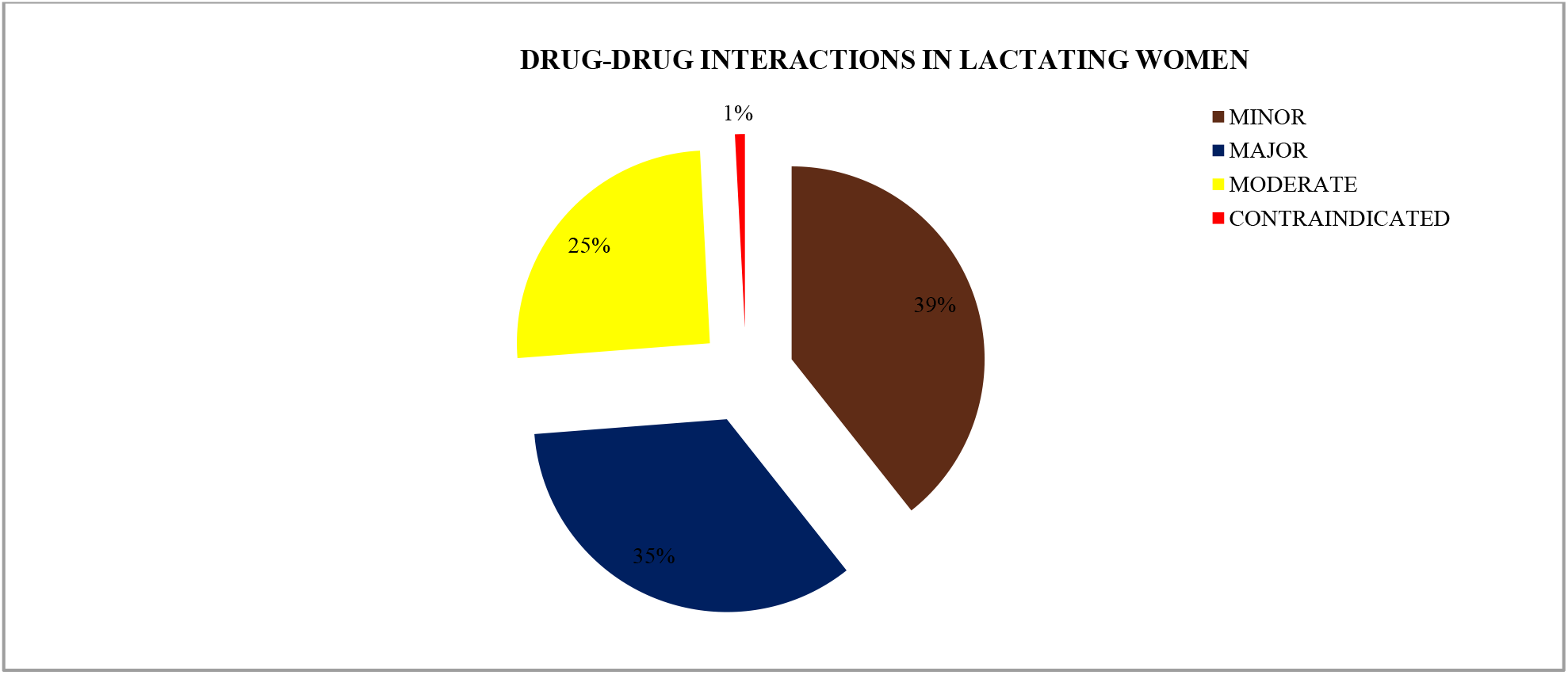
DDI in Lactating women

### Number of DDI’s distributed on the Basis of Per Prescription

A total of 206 prescriptions result were also calculated based on DDI’s present in each prescription. 62 (30.09%) prescriptions contained only 1 DDI, 25 (12.13%) prescriptions with two DDI’s, 17(8.25%) prescriptions contained three DDI’s, 14 (6.80%) prescriptions with four DDI’s, and 13 (6.31%) prescriptions with more than 5 (>5) DDI’s. Only 82 (39.8%) prescriptions were found to have no interaction.

**Table 10.** Number of Prescriptions and Percentage

| **<u>Number of DDI</u>** | **<u>Number of Prescription</u>** | **<u>Percentage</u>** |
| --- | --- | --- |
| **NIL** | 82 | 39.8% |
| **1** | 62 | 30.09% |
| **2** | 25 | 12.13% |
| **3** | 17 | 8.25% |
| **4** | 14 | 6.80% |
| **>5** | 13 | 6.31% |
| **N** | **206** |  |

**Figure 9.**
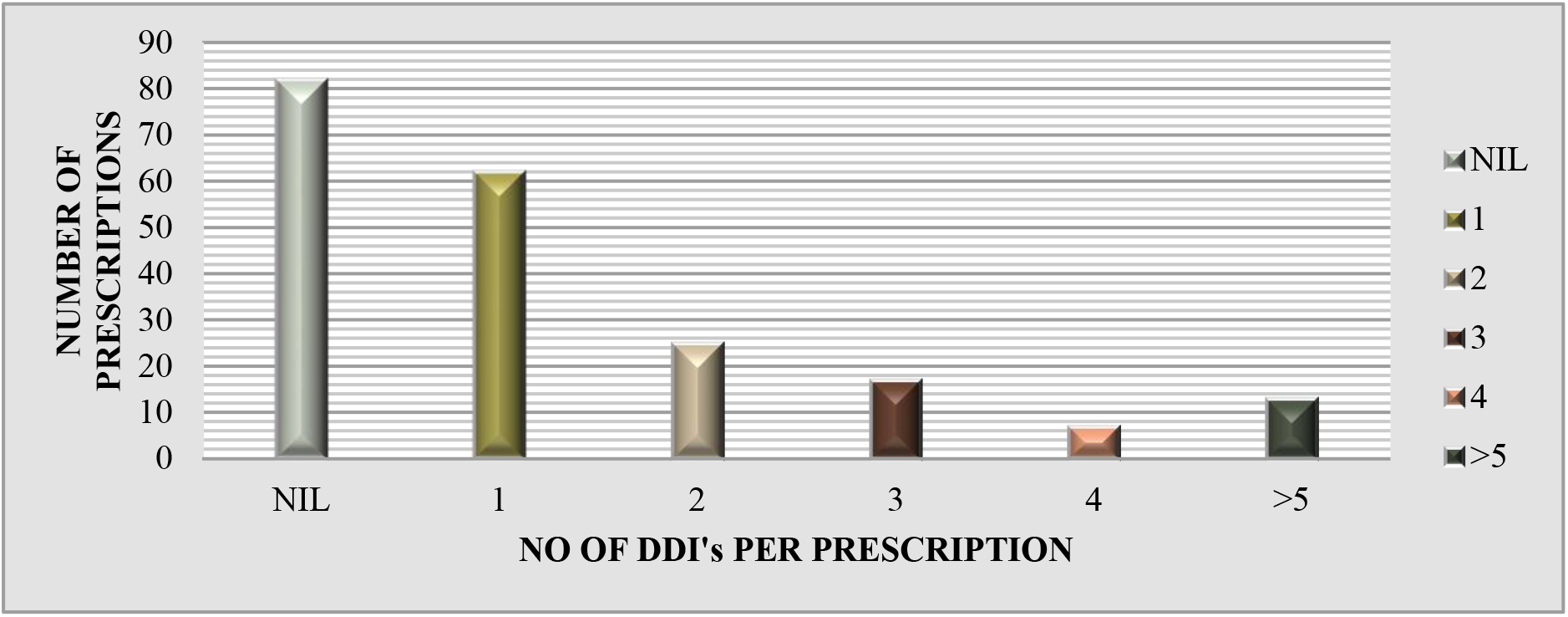
Number of DDI per prescription

### Distribution of DDI’s on the Basis of Age

Age wise DDI’s were distributed among total 214 interactions. Women of age group <20 were found with 27 (12.62%) interactions, 20-25 age group with 90 (42.06%) interactions, 26-30 age group with 77 (35.98%) interactions, 31-35 age group with 14 (6.54%) interactions, 36-40 age group with 4 (1.87%) interactions and > 40 age group with 2 (0.93%) interactions.

**Table 11.** Age wise distribution

| PATIENT'S AGE GROUP | <u>NUMBER OF DDIs</u> | Percentage of Total DDIs Founds |
| --- | --- | --- |
| <20 | 27 | 12.62% |
| 20-25 | 90 | 42.06% |
| 26-30 | 77 | 35.98% |
| 31-35 | 14 | 6.54% |
| 36-40 | 4 | 1.87% |
| >40 | 2 | 0.93% |
| **N** | **214** |  |

**Figure 10.**
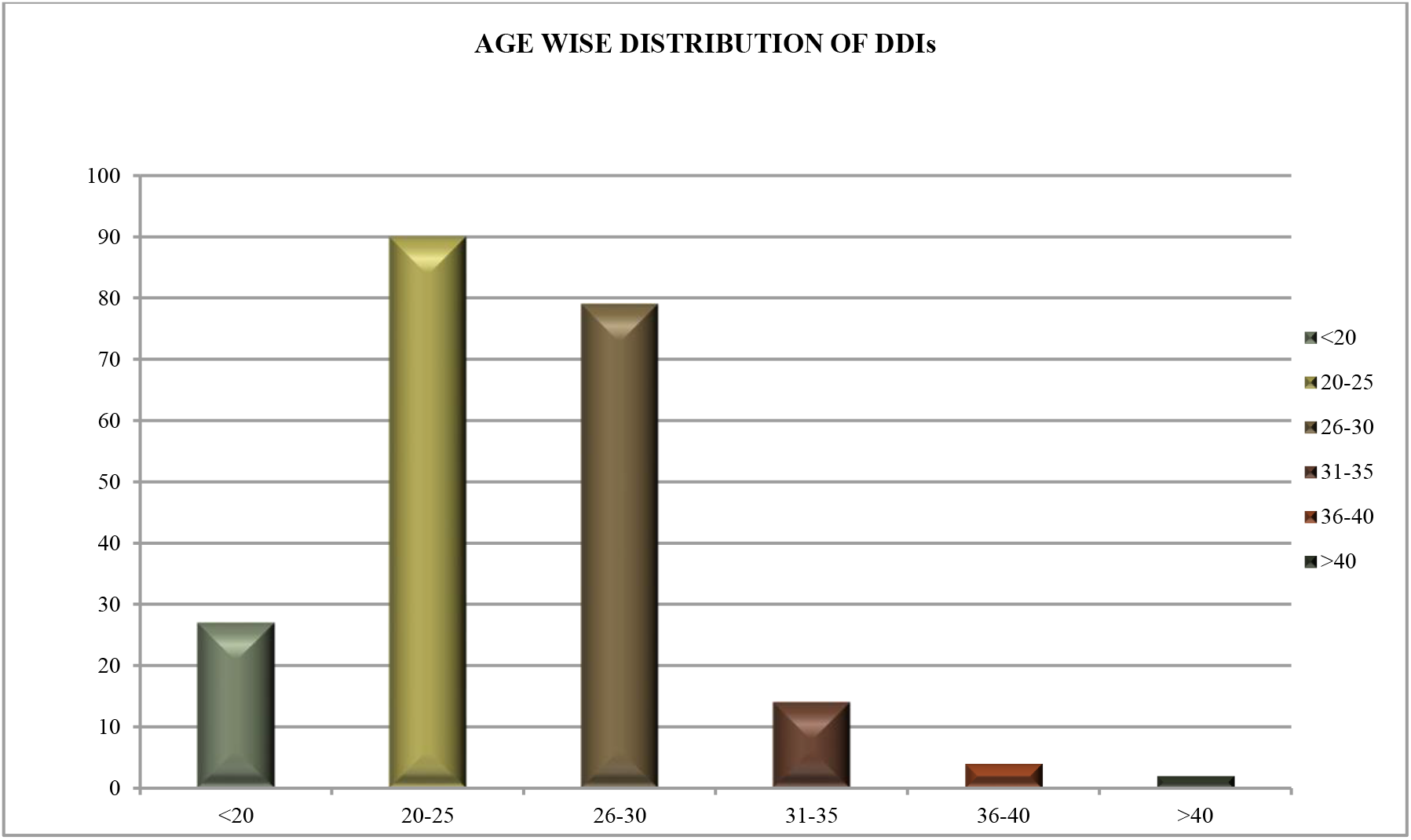
Distribution of DDI, s on the basis on age

### Pharmacokinetic Interactions

Drug-Drug Interactions in each prescription were also evaluated based on their pharmacokinetics and pharmacodynamics nature. Overall pharmacokinetic interactions were 89 out of which 33 (37.08%) were affecting the absorption of the drug, 3 (3.37%) were affecting the distribution, 24 (26.97%) were affecting the metabolism, and 29 (32.58%) affected the excretion of the drug.

**Table 12.** Pharmacokinetic interactions

| **<u>PHARMACOKINETIC</u>**<br>**<u>(PK)</u>**<br>**<u>INTERACTIONS</u>** | **PHASE Of PK** | **<u>NUMBER</u>**<br>**<u>Of DDIs</u>** | **<u>PERCENTAGE OF PK TYPE</u>**<br>**<u>DDIs</u>** |
| --- | --- | --- | --- |
|  | **Absorption** | 33 | 37.08% |
|  | **Distribution** | 3 | 3.37% |
|  | **Metabolic** | 24 | 26.97% |
|  | **Excretion** | 29 | 32.58% |
| **N** |  | 89 |  |

**Figure 11.**
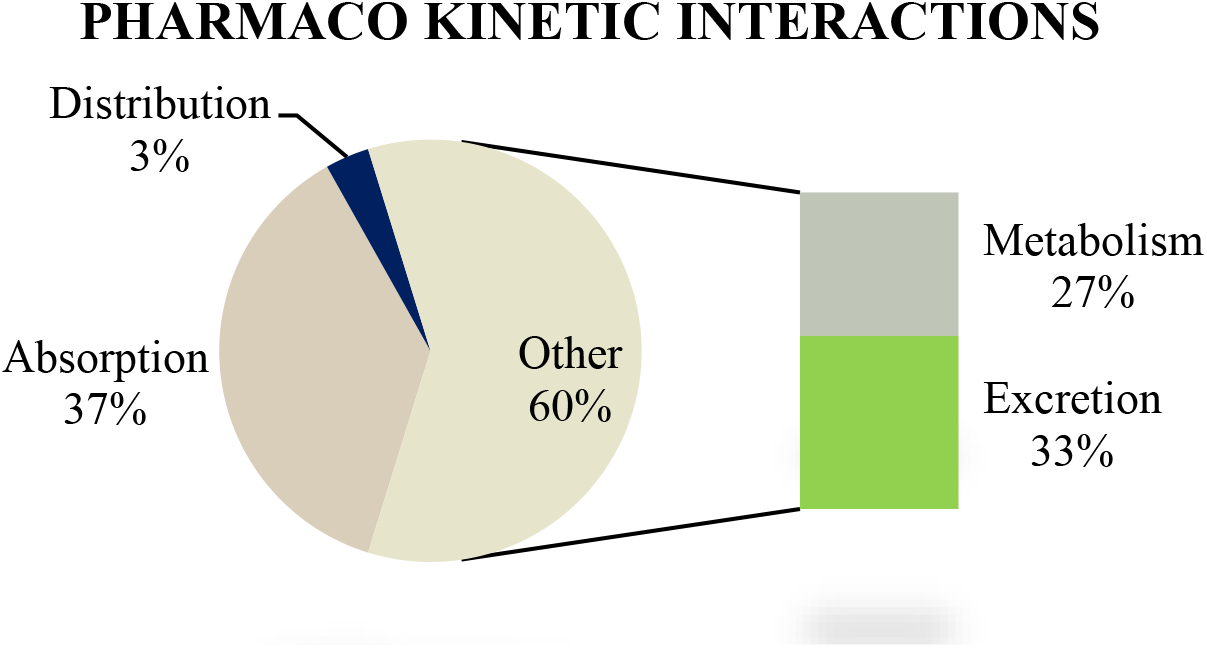
Pharmacokinetic interactions

#### Pharmacodynamics Interactions

Pharmacodynamics interactions evaluated in the prescriptions were 125. Out of which 37 (29.6%) were additive, 23 (18.4%) were synergistic and 65 (52%) were antagonistic pharmacodynamics interactions.

**Table 13.** Pharmaco-dynamic interactions

| **<u>PHARMACODYNAMIC</u>**<br>**<u>(PD)</u>**<br>**<u>INTERACTIONS</u>** | **<u>TYPE OF PD</u>**<br>**<u>INTERACTIONS</u>** | **<u>NO. OF</u>**<br>**<u>INTERACTIONS</u>** | **<u>PERCENTAGE</u>** |
| --- | --- | --- | --- |
|  | - |  |  |
|  | ADDITIVE | 37 | 29.6% |
|  | SYNERGISTIC | 23 | 18.4% |
|  | ANTAGONISTIC | 65 | 52% |
| **N** |  | **125** |  |

**Figure 12.**
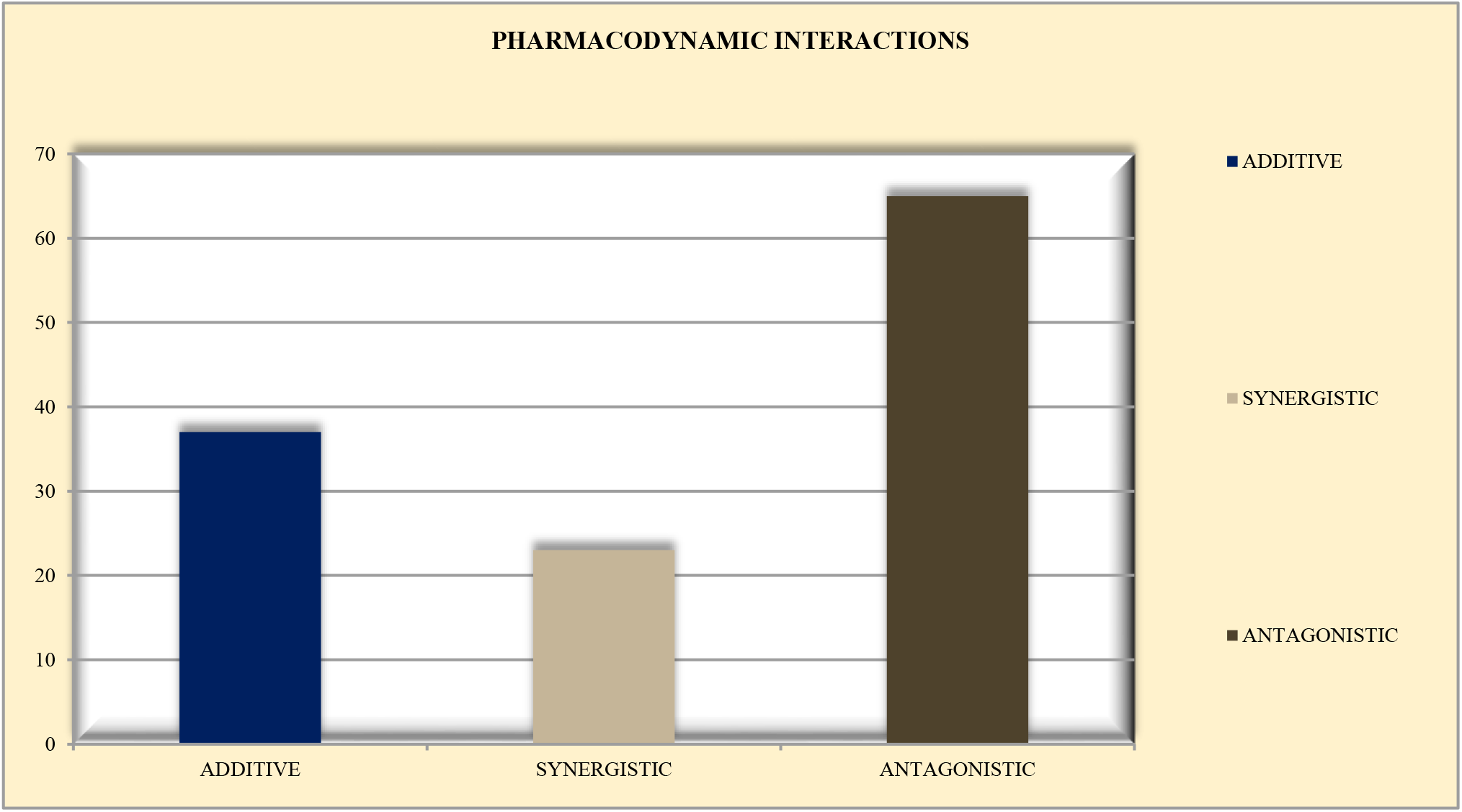
Pharmacodynamic interactions

## Discussion

Based on subgroups data was evaluated for pregnant and lactating women for the possible Drug-drug interactions and contraindications. 128 (62.0%) of the women involved in their research were pregnant women and 78 (38.0%) of them were lactating women.

206 cases were evaluated, and categorization based on the age groups was done to the study subgroups i.e. pregnant and lactating or breast-feeding women. Age groups included in the study were < 20 years, 20-25 years,26-30 years, 31-35 years, 36-40 years, and >40 years. The maximum number of females involved in the study were of age group 26-30 years i.e 91 and were 44.17% of all the age groups involved in the study. And > 40 years of age group contained only 1 woman i.e 0.48% of the 206 women subjects.

About 831 drugs were prescribed in 206 prescriptions collected, including post-partum prescriptions, pre-operative prescriptions, and prescriptions of lactating or breastfeeding women. Drugs prescribed for medication therapy in evaluated were antibiotics, NSAIDs, PPIs, Vitamin, minerals, supplements, iron, and folic acid, antifungals, antifibrinolytic and anticoagulants, antacids, antihypertensive and anti-angina agents, was bile acid, were anti-emetics, herbal drugs, corticosteroids, antispasmodic, hormones and enzymes, analgesics and antipyretics, opioids and benzodiazepine and miscellaneous (including intravenous infusions and drips, etc.). Most prescribed drugs in both subgroups were antibiotics i.e. cephalosporin, penicillin-like antibiotics, etc. and the total antibiotics prescribed were 213 which is 25.63% of the total medications prescribed and after antibiotics, another highly prescribed number of drugs were Vitamin, minerals, supplements, iron, and folic acid, which were 198 and about 23.82 % of the total prescribed medications. Analgesics and antipyretics prescribed were 81 which is 9.74 % of the total prescribed medications. The number of hormones and enzymes prescribed was comparatively less while comparing with analgesics and antipyretics. 1.68% of herbal drugs were also prescribed in the prescriptions and cranberry exact was the most prescribed herbal drug in pregnant and lactating women. As opioids and benzodiazepines are not given mostly in pregnancy and lactation but 0.96% of opioids were also prescribed in some prescriptions and one of the less prescribed classes of drugs found among prescriptions evaluated. Along with anti-hypertensive and anti-angina 0.96%, antacids 0.84%, and bile acids being the least prescribed drugs with 0.12% prescribing rate in the evaluated prescriptions.

Total cases were 206. And the number of prescribed drugs are 2, 3, 4, 5, 6, 7 and more than 7. This shows that multiple drugs are being prescribed in the pregnancy and lactation which is a very serious consideration, as it may lead to medication errors and ultimately leading to DDI’s. Most of the prescriptions contained 3 drugs i.e. 23.78% of the total prescribed medications. On the other hand 2^nd^ most prescribed number of drugs is 5 and 6drugs in the single prescription i.e. 16.01% and 16.99% of the total prescriptions collected. Prescriptions containing more than 7 drugs i.e, 6.31% prescriptions are mostly those which were collected for the pre-operative and post-operative patients.

Pregnant and lactating women were evaluated for possible Drug-Drug Interactions. In pregnant women number of interactions found was 91 (42.52%) while in lactating women number of interactions found were 123 (57.48%). Total contraindicated drugs prescribed in the prescriptions of pregnant and lactating women were 4 which was 0.48% of the total drugs prescribed from which 3 (75%) were found in prescriptions of pregnant women and 1 (25%) was found in the prescription of a lactating woman.

In pregnant women, out of 91 interactions found based on FDA risk categorization, the interactions were categorized as major, minor and moderate severity and drugs that were belonging to Category X of FDA risk categorization and hence placed in contraindicated drugs. Interactions of moderate severity i.e. 38.46% were most found interactions among the pregnant women and the contraindicated drugs prescribed were least found i.e. 4% evaluated according to FDA categorization of the drugs according to severity and risks.

123 drug-drug interactions were found in the lactating women were of a minor, major and moderate severity. Most repeated interactions in pregnant women were of minor severity and major severity i.e. 39.02% and 34.15% respectively. The least number was of the contraindicated drug which was about 1% of the total interactions in the lactating women and 15% of all the interactions found in both pregnant and lactating women.

From the total of 206 prescriptions results were also calculated based on DDI’s present in each prescription i.e. 1, 2, 3, 4, and >5 DDI’s per prescription. The highest number of DDI’s i.e., more than 5 were found in 6.31% of the prescriptions while in 39.8% of prescriptions, no DDI was found. 30.09% of the prescriptions contained a single DDI.

Age-wise DDI’s were distributed among a total of 214 interactions. Prescriptions of women of age group <20, 20-25, 26-30, 30-35, 36-40, and >40 were evaluated for interactions based on age groups, to know which age group has the highest prevalence of the DDI’s. From the results, it was evaluated that Age group 20-25 has the maximum number of interactions found in the prescriptions i.e., 41.67% and after those 26-30 years age group has 36.57% interactions in the evaluated prescription. Least percentage of interactions found in the age group >40 i.e. 0.92% which is due to the least number of the prescriptions available to be evaluated.

Drug-Drug Interactions in each prescription were also evaluated based on their pharmacokinetics and pharmacodynamics nature. Overall pharmacokinetic interactions were 91 out of which 33 (37.93%) were affecting the absorption of the drug, 3 (3.44%) were affecting the distribution, 24 (27.58%) were affecting the metabolism, and 31 (31.03%) affected the excretion of the drug. Most occurring pharmacodynamics interactions were affecting the absorption of the drug in the percentage of 37.93% and least affected was the distribution of drug i.e., 3.44%. In pregnancy, although the pharmacokinetic parameters change for the women’s body which affects the ADME (Absorption, Distribution, Metabolism, and Excretion) of the drugs. Hence, these interactions are showing the effect of one drug on the other to alter its pharmacokinetics.

Pharmacodynamics interactions evaluated in the prescriptions were 123. They were categorized as an additive, synergistic and antagonistic. Most found pharmacodynamics interactions were 56.92% of the additive type and least found were 13.84% of the synergistic interactions.

All these results have shown the evaluation of Drug-Drug Interactions from the different aspects in the prescriptions of the evaluated subgroups i.e. pregnant and lactating women whether it was based on age groups or the basis of pharmacokinetics and pharmacodynamics. All these results added specificity to data and created a clear picture of DDI’s by categorizing them into different parts.

## Conclusions

From our study we demonstrated a significant number of drug-drug interactions in both pregnant and lactating mothers due to the complexity of prescribed pharmacotherapy and multiple drug prescribing. Most of the interactions found in our prescriptions had minor and major levels of severity. A small percentage of contraindicated drugs were also found for both pregnant and lactating women which could be easily substituted with safer medicine. Drug-drug interaction prevalence is directly related to the number of prescribed medicines and medication errors. This study also emphasizes the role of pharmacists to check and review the prescribed medicines closely to prevent detrimental drug-drug interactions and their unfavorable impacts on both mother and child.

## Data Availability

All data produced in the present work are contained in the manuscript.

